# Effect of Therapeutic Inertia on Treatment Outcomes in Type 2 Diabetes Patients at a Tertiary Care Hospital in Southern Ethiopia: A retrospective Cohort Study

**DOI:** 10.1101/2025.06.01.25328748

**Authors:** Dawit Alemu Lemma, Lijalem Abera Tema, Muluken Berhanu Mena, Hailu Chare Koyra

**Affiliations:** wachemo University College of medicine and health science, department of pharmacy, hosaina Ethiopia, Wolaita Sodo University, College of health sciences and Medicine; Department of Internal Medicine, Wolaita Sodo, Ethiopia; Wolaita Sodo University, College of health sciences and Medicine; Department of Pharmacy, Wolaita Sodo, Ethiopia

**Keywords:** Therapeutic inertia, treatment intensification, type 2 diabetes, treatment outcomes

## Abstract

**Aims:** Therapeutic inertia is the failure to initiate or intensify treatment when clinically indicated, remains a major challenge to optimal glycemic control in patients with type 2 diabetes. This retrospective cohort study investigated the effect of therapeutic inertia on treatment outcomes in patients with type 2 diabetes receiving care at a tertiary hospital in southern Ethiopia.

**Materials and methods:** A retrospective cohort study was conducted among 159 adult ambulatory type 2 diabetic patients between June 2020-2023. We collected the data from medical records and we used EpiData version 4.6 for data entry and SPSS version 25 for analysis. The independent sample t-test, fisher’s exact test and chi-square test were used for data analysis as appropriate. To assess the effect of therapeutic inertia on diabetic treatment outcomes, we applied a cox proportional hazard model. A p-value of less than 0.05 was considered statistically significant.

**Results:** In this study, we reviewed the medical records of type 2 diabetic patients. Poor treatment outcomes were common in exposed to therapeutic inertia group (68.63%).There was a statistically significant difference between groups in the type of physician managing the patients (p=0.01).furthermore, the presence of comorbidity (p=0.024), mean fasting plasma glucose level (p=0.01), neuropathy(p=0.02) and nephropathy (p=0.011) showed a significant association with therapeutic inertia. Therapeutic inertia exposed group was significantly associated with increased the risk of poor treatment outcome with adjusted HR of 1.927 (95% CI: 1.201-3.092, p = 0.007).

**Conclusions:** Our study showed therapeutic inertia has worsened diabetic treatment outcomes among type 2 diabetes patients. This underlines the Healthcare providers should prioritize proactive management, such as regular reassessment of treatment efficacy and prompt adjustment of therapies, to improve patient outcomes.

## Introduction

Diabetes mellitus (DM) describes a group of chronic metabolic disorders. The characteristic feature of DM is hyperglycemia, which can lead to long-term microvascular and neuropathic complications and is an important contributor in newly diagnosed adult blindness cases, end-stage renal disease, and non-traumatic lower limb amputations. DM additionally contributes to macrovascular complications like coronary artery disease, peripheral vascular disease, and stroke[1].

The vast majority of diabetic patients fall into one of two broad categories: type 2 diabetes mellitus (T2DM), which is characterized by the presence of insulin resistance with an inadequate compensatory increase in insulin secretion, or type 1 diabetes mellitus, which is caused by an absolute or nearly absolute deficiency of insulin [2]. The prevalence of type 2 diabetes (T2DM) is expected to reach 7079 people per 100,000 by 2030, with a disability-adjusted life years (DALYs) change of 1.86 percent yearly, demonstrating an ongoing rise in cases across all regions of the world and adding to the growing global burden of diabetes mellitus. Rising prevalence patterns in low-income nations are worrying [3, 4].

The growing prevalence of non-communicable diseases like diabetes is currently a problem in Ethiopia[5]. According to the world health organization (WHO), 3.2 % of Ethiopians has diabetes mellitus. According to other study, the prevalence of diabetes mellitus in Ethiopia ranges between 0.5 to 6.5 %[6].

Achieving blood glucose levels within the target range is widely recognized as an important strategy for reducing the risk of the development and progression of type 2 diabetes (T2DM)related complications[7, 8]. The cornerstone of the current strategy for managing T2DM is the United Kingdom Prospective Diabetes Study (UKPDS), which shows that tighter glycemic control has been associated to a better treatment outcome and fewer complications [9, 10].

The American Association of Diabetes (ADA) and the European Association for the Study of Diabetes (EASD) both agree that the target value of HbA1C that we should strive for in treating the majority of patients should be ≤7 % [10, 11]. For some people, a tighter target of 6.5% would be appropriate if it could be achieved without resulting in significant adverse effects, including hypoglycemia. These patients may be younger, have a longer expected life expectancy, have had diabetes for a shorter time, be receiving only metformin or lifestyle changes, or have no significant comorbidities [12]. Less stringent targets, such as 8%, may be appropriate for older individuals, who have had diabetes for a long time, have experienced severe hypoglycemia in the past, or have numerous serious comorbidities [12]. To prevent the onset and progression of chronic complications, tight glycemic control is essential. Stepwise intensification of antidiabetic medication is also widely recommended [13, 14].

Regrettably, it is very challenging to meet the objectives from the guidelines in routine clinical practice, despite individualizing treatment and clear recommendations. There are many reasons for this situation, and one of them can be covered by the term “therapeutic inertia” [15].

Therapeutic inertia is the delay in initiating or intensifying therapy when it is appropriate to do so; despite the glycemic target goals are not being met [16, 17]. The word “inertia” generally has a negative connotation, suggesting that the physician is entirely responsible for not giving treatments appropriately or as intensely as they should be, in order to ensure that patients reach guideline recommended goals. Therapeutic inertia in T2DM is a concern, along with anti-diabetic medication and other therapy aiming to lower cardiovascular risk (antihypertensive drugs, lipid lowering agents, antiplatelet therapies)[17].

Factors that may contribute to therapeutic inertia can be categorized into three groups. The first group of factors is depends on the physician. The second category consists of patient-related factors. The third group focuses on the healthcare system [18].

When considered collectively, a variety of factors at the patient, provider, and health-system levels have an impact on the capacity to provide care for diabetic patients. All these contribute to the problem of therapeutic inertia, which has a major effects on treatment outcomes for diabetic patients[18]. Despite the growing recognition of therapeutic inertia as a major barring factor for optimal glycemic controls. There is limited evidence from low-resource settings, especially the sub-Saharan African region, regarding how therapeutic inertia comes to play and how it affects treatment outcomes, most available studies have been conducted in high-income countries, while very little attention has been paid to the contextual challenges faced in Ethiopian healthcare settings. Therefore, this study would be an attempt to close this crucial gap existing in the local as well as in the global literature by analyzing the effect of therapeutic inertia on treatment outcome among patients with type II diabetes attending a tertiary hospital situated in southern Ethiopia.

## Materials and methods

The study was carried out in Ethiopia, at the Wolaita Sodo University Comprehensive Specialized Hospital which is approximately 329 kilometers away from Addis Ababa, the capital city of Ethiopia. The aim of the study was to determine therapeutic inertia and its effect on diabetes treatment outcomes.

From 18 July 2023, data for this retrospective study were accessed continuously for thirty days. Authors never had access to any information that could identify individual participants during or after data collection since all data were fully anonym zed before analysis. The study involved adults aged above 18 years with complete medical records, at least 3 years of treatment duration and have uncontrolled diabetes. Patients, who were pregnant, had pre-existing complications, comorbidities with CCI >3, were above 65 years of age, had substance use or psychiatric disorders, were treated with insulin monotherapy, or had complications at the time of diagnosis were excluded. Data collection was retrospective and aimed at capturing, through a structured format, demographic information, laboratory results (HbA1c, lipid profiles, and renal functions), treatment histories, and diabetes-associated complications which were diagnosed by physicians. The burden of comorbid conditions was identified with the Charlson Comorbidity Index which is used to determine prognosis mortality risks.

The sample size was determined using a G*Power software, a sample size of 159 patients was selected based on a significance level of 5%, power of 80%, and previously estimated therapeutic inertia prevalence of 38.35%[19].With the additional 10% contingency. From this 332 medical records were excluded from the study and 460 records met the eligibility criteria. For the non-therapeutic inertia (non-exposed) group 57 records fulfilled the inclusion criteria. For the therapeutic inertia (exposed group), we used a systematic random sampling technique using a total of 102 patient records were gathered. In our study, the exposed to non-exposed ratio was nearly 2:1(1.79:1).

For this study, therapeutic inertia was characterized by the proportion of patients with no observable actions in their index date; Patients were followed every three months up to the 36 months which is the end of our follow-up. An observable action was defined as either: receiving a prescription within three months for a new category of glucose-lowering medications or increase the doses in addition to the existing baseline medication regimen, or not receiving a new prescription but maintaining a measured glycemic value within the target range.

We selected a glycemic target of 80–130 mg/dl of mean FBs in this research since individualized targets are not available in our source data and after applying our eligibility criteria, which excludes the majority of patients in less and stricter targets.

### Statistical methods

The primary dependent variable was treatment outcome, while the key independent variable was therapeutic inertia status. Additional factors analyzed included patient demographics (age, sex, residency), clinical factors (diabetes duration, medication history, glycemic levels), and physician-related variables (qualification, visit frequency). Therapeutic inertia was defined according to the ADA 2019 guidelines, which outline when therapy intensification is required. Outcomes such as hospitalization, mortality, and glycemic control were monitored at baseline and at three-month intervals.

The statistical package for social sciences (SPSS) version 25 was used to analyze the data once it was entered into Epi-Data V.6. Results were presented as descriptive statistics (frequencies, percentages, means, median and standard deviations), depending on the type of variables and the normality of the distribution.

Therapeutic inertia group characteristics were compared to non-inertia group characteristics using an independent t-test for continuous variables based on the distribution of data and a chi-square or Fisher’s exact test for categorical data. P values of less than 0.05 were considered statistically significant. The Cox proportional hazards model was used to determine the effect of therapeutic inertia diabetic treatment outcomes and adjustments for confound variables were also made.

Figure 1 shows the patient screening process. A total of 792 patients attended the clinic in 2020, in which 460 of them had blood glucose levels higher than targets. After excluding 301 patients who met the exclusion criteria, 159 remained for the study. These were then categorized into two groups according to whether or not they had experienced therapeutic inertia: 102, while 57 did not.

**Figure 1:**
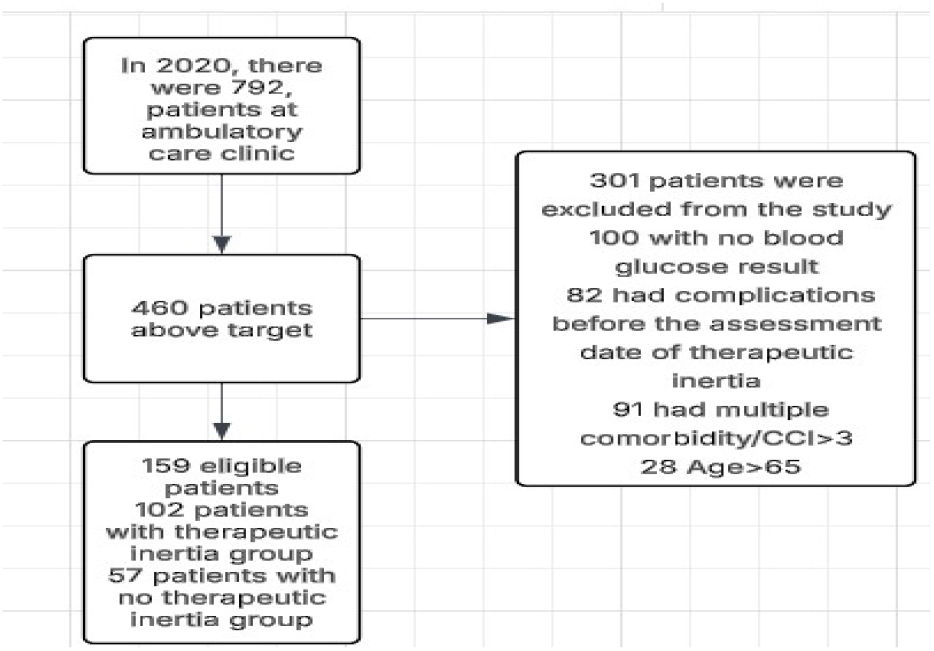
Flow diagram reporting the number of patients excluded at each stage of recruitment.

## Results

### Population demographics and baseline characteristics

In this study we assessed medical records of 159 type 2 diabetic patients, 6.86% were lost to follow-up in the exposed group and 1.75% in the non-exposed group and 7.1% in the exposed group completed the study without treatment intensification. As shown in Table 1 Based on gender 61.8% of men and 38.24% of females were in the therapeutic inertia exposed group. Participants who lived in urban areas (69.62%) were higher in the Therapeutic inertia exposed group compared to ruler areas (30.4%) and 16.7% had insurance coverage in the therapeutic inertia exposed group. Age distribution showed Participants in the age group of 40-49 were in a more therapeutic inertia group at 42.2% and relatively low in those less than 30 years of age, at 2.9%, and when treated by general practitioners (89.22%) compared to specialists (2.94%) in therapeutic inertia group.

**Table 1:** Socio-demographic and clinical characteristics of study participants at WSUCSH (n=159)

| Characters | Category | Therapeutic inertia<br>Frequency (%)<br>102(64.2) | Therapeutic inertia<br>No, frequency (%)<br>57(35.8) |
| --- | --- | --- | --- |
| Age | <30 | 3(2.9) | 3(5.3) |
|  | 30-39 | 8(7.84) | 7(12.3) |
|  | 40-49 | 39(38.24) | 21(36.84) |
|  | 50-59 | 31(30.4) | 13(22.81) |
|  | ≥60 | 21(20.6) | 13(22.81) |
| Gender | Male | 63(61.8) | 31(54.4) |
|  | Female | 39(38.24) | 26(45.61) |
| Duration of T2DM | <10 | 98(96.08) | 54(94.76) |
|  | ≥10 | 4(3.92) | 3(5.26) |
| Insurance coverage | Yes | 17(16.67) | 12(21.1) |
|  | No | 85(83.3) | 45(78.95) |
| Place of residence | Urban | 71(69.62) | 44(77.2) |
|  | Rural | 31(30.4) | 13(22.81) |
| Type of physician | General practitioners | 91(89.21) | 28(49.12) |
|  | Residents | 8(7.84) | 4(7.02) |
|  | Specialists | 3(2.94) | 25(43.86) |
| Antihypertensive medications | ACEIs/ARBs |  |  |
|  | Yes | 40(39.22) | 40(39.22) |
|  | No | 62(60.78) | 33(57.895) |
|  | CCBs |  |  |
|  | Yes | 32(31.37) | 16(28.07) |
|  | No | 70(68.63) | 41(71.93) |
| Comorbidity | Yes | 72(70.6) | 30(52.6) |
|  | No | 30(29.41) | 27(51.9) |
| Antiplatelet/anticoagulant | Yes | 23(22.6) | 34(59.65) |
|  | No | 79(77.45) | 44(40.36) |
| Statin use | Yes | 68(66.67) | 29(50.88) |
|  | No | 34(33.33) | 28(49.12) |

### Baseline socio-demographic characteristics of Participants compared between therapeutic inertia and no therapeutic inertia group

We compared therapeutic inertia (exposed) group baseline characteristics to non-inertia group characteristics using an independent t-test for continuous variables based on the distribution of data and a chi-square or Fisher’s exact test for categorical data. As demonstrated in Table 2, there were no statistically significant differences in gender distributions (p=0.364) and age (p=0.708). There was no significant difference between the groups in terms of T2DM duration or insurance coverage (p=0.702 and p=0.492, respectively). Statistically significant difference was shown between types of physician (p=0.001).

**Table 2:** Baseline socio-demographic characteristics of patients compared between therapeutic inertia and no therapeutic inertia group at WSUCSH between Jun 2020-2023(n=159).

| Variables | Category | Total | Therapeutic inertia | No-Therapeutic inertia | Variables |
| --- | --- | --- | --- | --- | --- |
| Gender | Male | 94 | 63 | 31 | 0.364 |
|  | Female | 65 | 39 | 26 |  |
| Age | <30 | 6 | 3 | 3 | 0.708 |
|  | 30-39 | 15 | 8 | 7 |  |
|  | 40-49 | 60 | 39 |  |  |
|  | 50-59 | 44 | 31 | 13 |  |
|  | ≥60 | 34 | 21 | 13 |  |
| Duration of T2DM | <10 | 152 | 98 | 54 | 0.702** |
|  | ≥10 | 7 | 4 | 3 |  |
| Insurance coverage | Yes | 29 | 17 | 12 | 0.492 |
|  | No | 130 | 85 | 45 |  |
| Type of physician | General practitioners | 119 | 91 | 28 | 0.001* |
|  | Residents | 12 | 8 | 4 |  |
|  | Specialists | 28 | 3 | 25 |  |

Frequency is used to characterize categorical variables. \*\* For categorical variables, fisher's exact test. \* p value <0.05.

### Baseline clinical characteristics of patients compared between therapeutic inertia and no therapeutic inertia group

As illustrated in Table 3 the presence of comorbidity (p=0.024) and mean fasting glucose level (p=0.001) showed a statistically significant difference between groups. Neuropathy and nephropathy were significantly associated (p=0.02, p=0.011) with therapeutic inertia. There was no statistically significant difference shown between the group for CCI score, systolic or diastolic blood pressure, and cardiovascular disease (p=0.933, p=0.093, p=0.400, p=0.723).

**Table 3:**
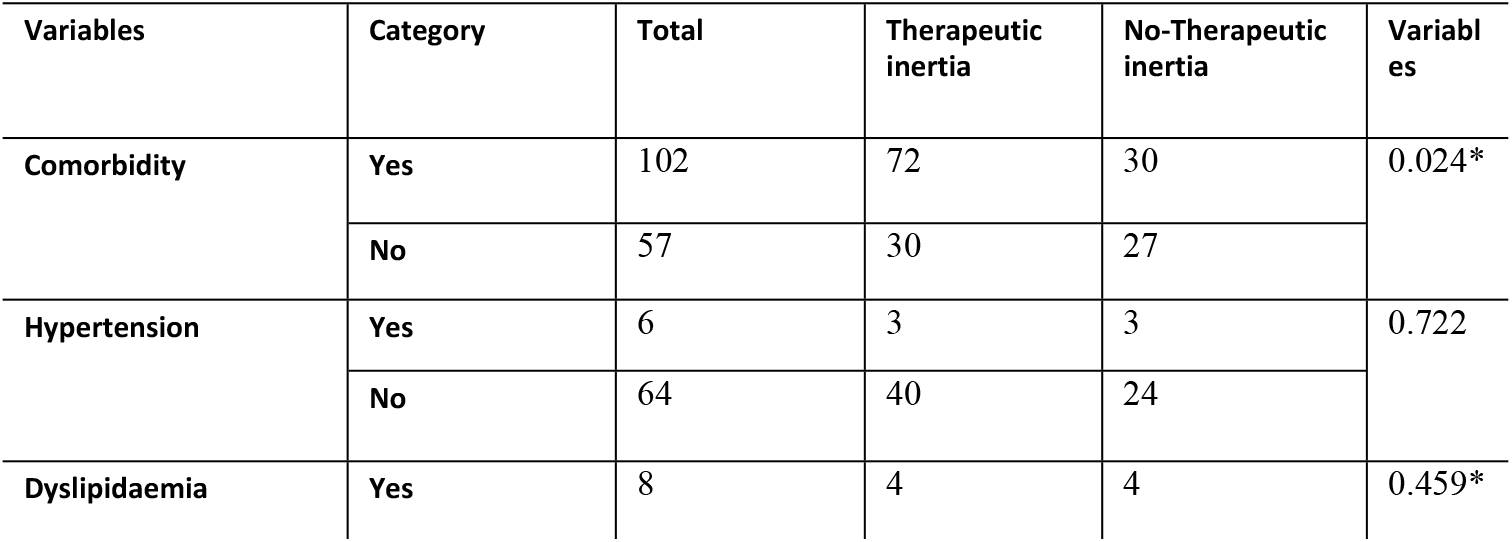
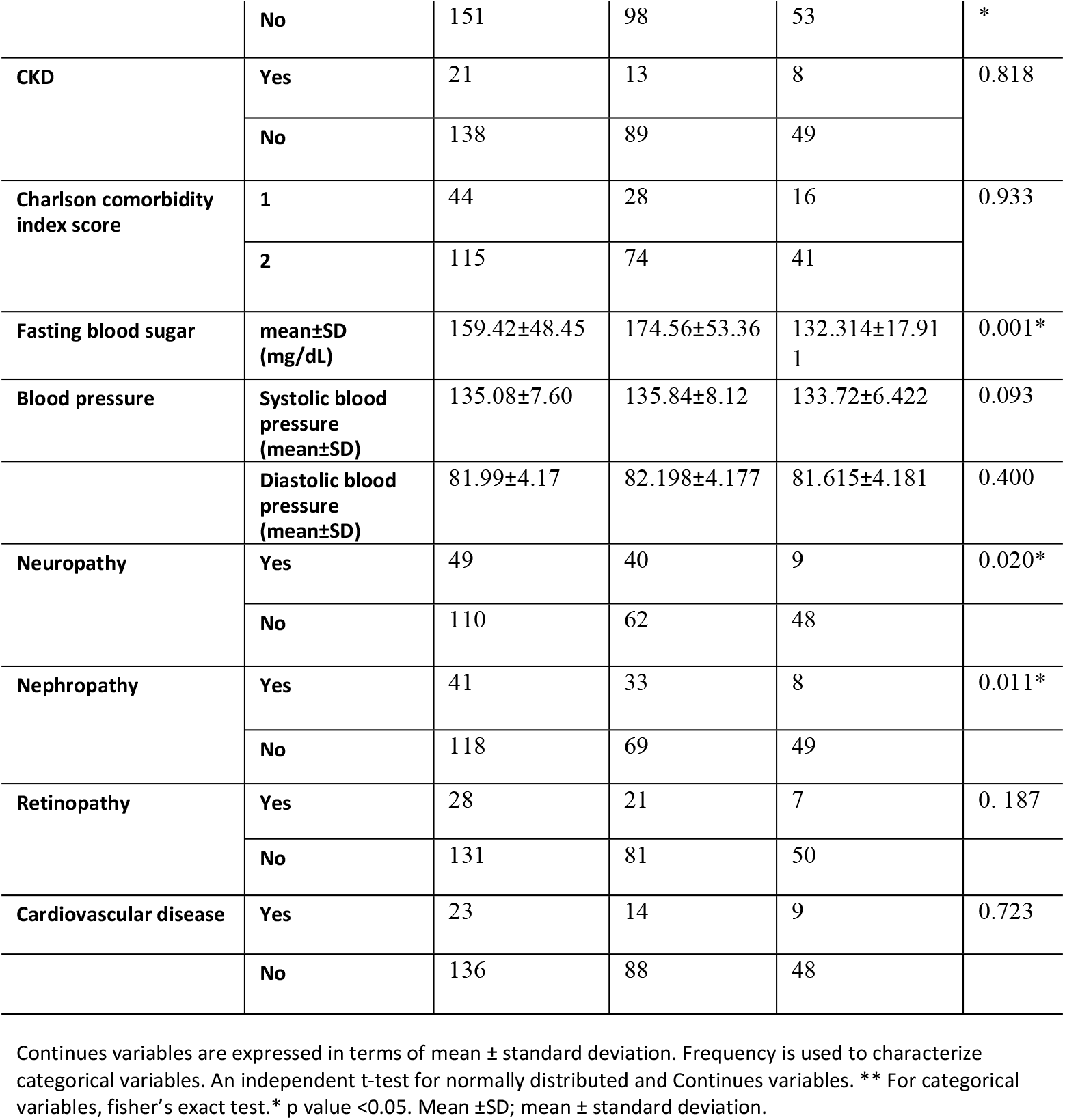
Baseline clinical characteristics of patients comparing between therapeutic inertia and no therapeutic inertia group at WSUCSH between Jun 2020-2023(n=159)

### Baseline treatment characteristics of patients compared between therapeutic inertia and no therapeutic inertia group

As shown in Table 4 there were no significant differences between groups in antidiabitic medications uses (P value =0.253).We assessed the use of antihypertensive and lipid lowering medications showed no significant difference in the utilization of ACEIs/ARBs (P value =0.722), CCBs (P value 0.664) or statin usages (P value =0.050).

**Table 4:**
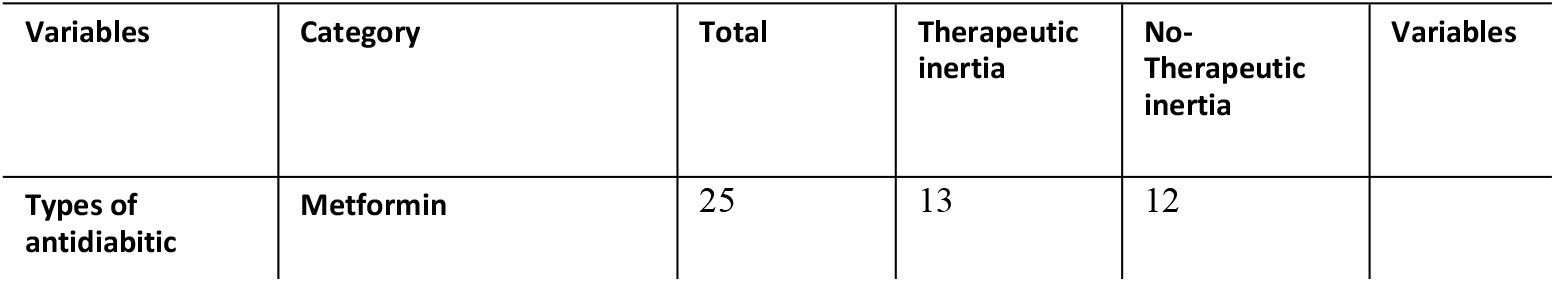
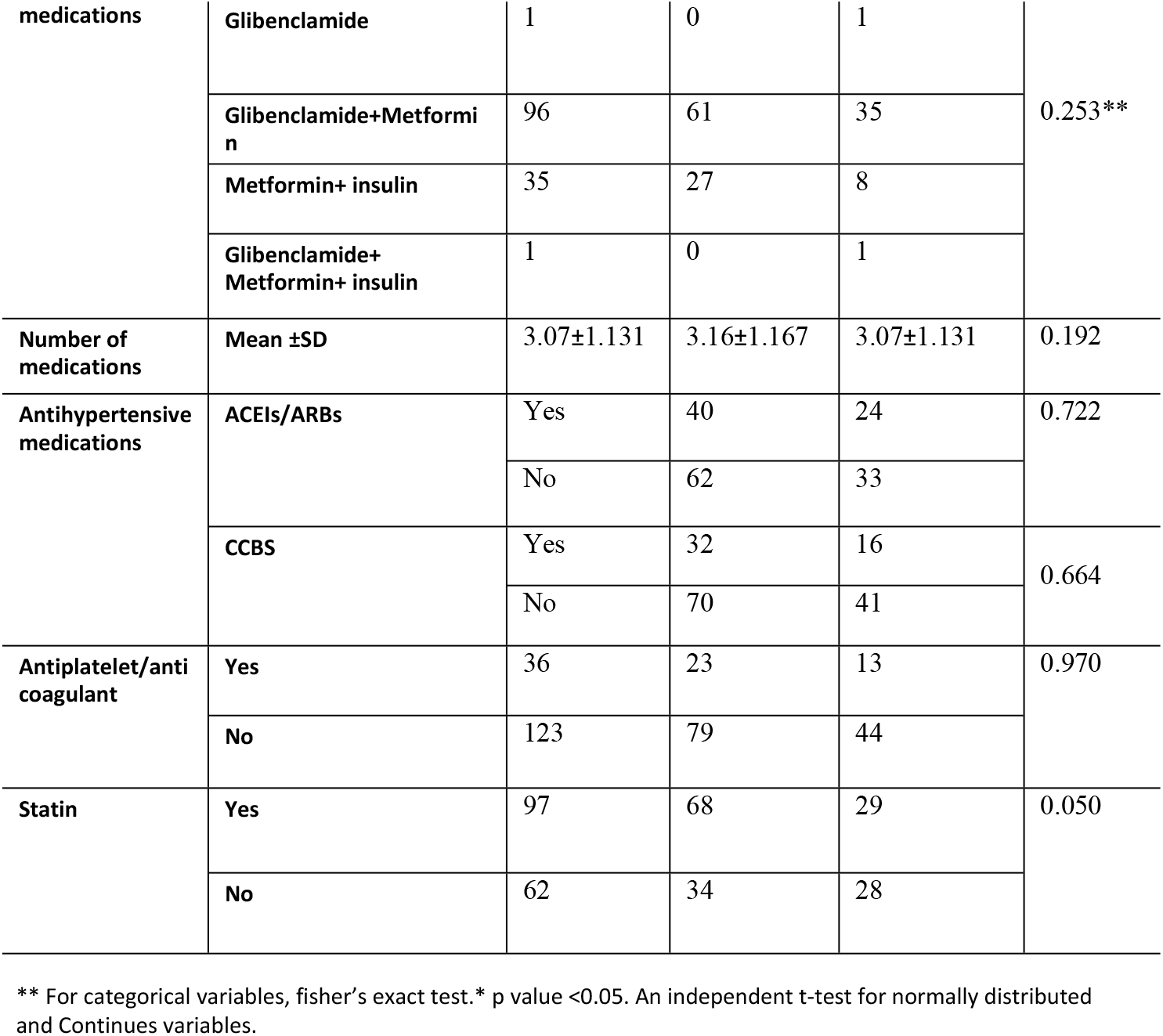
Treatment characteristics of patients comparing between therapeutic inertia and no therapeutic inertia group at WSUCSH between Jun 2020-2023(n=159).

### Cox regression analysis of the effects of therapeutic inertia on diabetic treatment outcome among type 2 diabetic patients

We conducted a bivariable Cox regression analysis to identify those variables candidates for multivariable analysis, as shown in Table 5 therapeutic inertia (exposed) was significantly associated with poor treatment outcome with CHR of 1.917 (95% CI :1.249-2.943, P = 0.003). Additionally, age categories, use of lipid lowering agent, number of medication used were candidates for multivariable analysis. In multivariable analysis among these factors, therapeutic inertia exposed group has a 92.7% higher risk of having poor treatment outcome compared to the non-exposed group. With an adjusted hazard ratio (AHR) of 1.927 (95% CI 1.201-3.092, p = 0.007).

**Table 5:** Multivariable cox regression on the effects of therapeutic inertia among type 2 diabetic patients at WSUCSH (n=159).

| Variables | Category | Diabetic treatment outcome |  | CHR(95% CI) | AHR(95% CI) | Variables |
| --- | --- | --- | --- | --- | --- | --- |
|  |  | Poor | Censored |  |  |  |
| Exposure status | Exposed | 70 | 32 | 1.917 (1.249-2.943) | 1.927 (1.201-3.092) | 0.007** |
|  | Non-exposed | 33 | 24 |  |  |  |
| Age | <30 | 3 | 3 |  |  |  |
|  | 30-39 | 8 | 7 | 3.527 (0.842-14.780) | 4.140 (0.804-21.305) | 0.505 |
|  | 40-49 | 44 | 16 | 2.65 (0.622-11.290) | 3.798 (0.848-17.023) | 0.089 |
|  | 50-59 | 26 | 18 | 2.994 (0.699-12.817) | 3.444 (0.744-15.930) | 0.081 |
|  | ≥60 | 22 | 12 | 3.92 (0.815-18.856) | 3.317 (0.734-14.999) | 0.114 |
| Use of lipid lowering agent | Yes | 62 | 35 | 1.379 (0.920-2.067) | 1.262 (0.787-2.024) | 0.335 |
|  | No | 41 | 21 |  |  |  |
| Number of medication | One | 10 | 2 |  |  | 0.240 |
|  | Two | 15 | 15 | 2.713 (0.594-12.397) | 2.555 (0.512-12.745) | 0.253 |
|  | Three | 53 | 22 | 1.436 (0.327-6.300) | 1.320 (0.284-6.143) | 0.724 |
|  | Four | 19 | 7 | 1.929 (0.468-7.948) | 1.658 (0.390-7.049) | 0.493 |
|  | Five | 4 | 5 | 1.867 (0.429-8.121) | 2.216 (0.490-10.029) | 0.302 |
|  | Others | 2 | 5 | 0.889 (0.161-4.901) | 0.712 (0.122-4.153) | 0.706 |

\*\* P value <0.05

## DISCUSSION

We conducted a study on therapeutic inertia and its effect on treatment outcomes among type 2 Diabetes Mellitus patients. Our findings determined the effect of therapeutic inertia on treatment outcomes of diabetic patients including, glycemic control, diabetic ketoacidosis (DKA), neuropathy, nephropathy, retinopathy, diabetic foot ulcer, metabolic control, cardiovascular diseases (CVD), death, referral, and hospitalization and also we compared therapeutic inertia effect between the two groups with poor and good treatment outcomes.

In our study, the average fasting blood sugar (FBS) level among patients with therapeutic inertia was 174.56 ± 53.36 mg/dL. In contrast, a study conducted on diabetic patients with therapeutic inertia in Cameroon reported an average FBS level of 200 ± 108 mg/dL[20]. The discrepancy in these results could be due to the fact that our study excluded patients who were only dependent on insulin. Both studies, however, indicated elevated FBS levels, specifically, our findings showed that patients experiencing therapeutic inertia had a mean FBS level of 174.56 ± 53.36 mg/dL. This suggests an association between delayed treatment intensification and higher FBS levels.

In our study, the types of physician involved in the treatment of type 2 diabetic patients showed that 75.5% were treated by general practitioners, 22.0% by residents, and 2.5% by specialists. A statistically significant difference was seen between the types of physicians and exposure status (p=0.001). In our study, 89.2% of patients treated by general practitioners were in the therapeutic inertia exposed group, while residents accounted for 7.8% and specialists for 2.9%. This result is in contrast to another study reporting that 32.2% of patients treated by general practitioners, 30.6% treated by residents, and 37.2% treated by specialists were in the therapeutic inertia exposed group[21]. The difference could be because most of the patients (43.4%) in the reference study were treated by specialists compared to ours.

The higher percentage of patients in therapeutic inertia group treated by general practitioners in our study suggests a potential role of physician specializations in influencing the timely intensifications of diabetic treatment. This shows the importance of targeted intervention and further training for general practitioners to address therapeutic inertia effectively.

In our study, there were significant differences between groups in the presence of comorbidities with 70.59% in therapeutic inertia exposed group and 29.41% in non-exposed group, (p=0.024). This result is consistent with a study done in Serbia where 76.8% of patients with comorbidities were in the therapeutic inertia group compared to 62.9% in a non-inertia group with a (p value=0.001) [22].

The current study identified a significant association between therapeutic inertia and nephropathy, with 32.35% in the therapeutic inertia group compared with 14.04% in the non-therapeutic inertia group (p-value = 0.011). Conversely, a study done in Taiwan reported slightly higher nephropathy cases in the non-therapeutic inertia group (10.8%) compared with the therapeutic inertia group (7.9%). Differences in findings may be due to our study which showed a higher percentage of patients experiencing therapeutic inertia (64.2%).In contrast, the reference study found a higher percentage of patients without therapeutic inertia (75.9%)[23].These differences could also be influenced by differences in the quality of care provided.

In our study a significant association between therapeutic inertia and neuropathy was identified, with 39.22% in therapeutic inertia group compared with 15.79% in the non-therapeutic inertia group (p-value = 0.020), similarly a study conducted in Cameroon reported 26% of participants had diabetic nephropathy[24].

In our study, therapeutic inertia was significantly associated with poor treatment outcomes with an adjusted hazard ratio (AHR) of 1.927 (95% CI 1.201-3.092, p = 0.007) compared to non-exposed individuals. Our result is in contrast to a study done in the US (AHR, 1.10; 95% CI, 1.03–1.07; P = 0.004) [25]. This discrepancy is most likely due to variations in patient demographics, healthcare professional practices, and healthcare infrastructure between our study and the reference study. However both study reported that delayed treatment intensification was associated with poor treatment outcomes.

In another study, both our study and the study from Thailand gave important results on the effect of therapeutic inertia on diabetic treatment outcomes among patients with diabetes. In our study, therapeutic inertia was significantly associated with an increased risk of poor treatment outcomes, with an adjusted hazard ratio (HR) of 1.927 (95% CI 1.201-3.092, p = 0.007). This result is consistent with the Thailand study, which found an adjusted HR of 1.51 and a substantial increase in the risk of poor treatment outcomes associated with therapeutic inertia [23].

This study showed therapeutic inertia was associated with an increase in the risk of poor treatment outcomes (adjusted HR: 1.927, 95% CI, 1.201-3.092, p = 0.007).In contrast, a US study reported a 19% increase in overall poor diabetic treatment outcomes for patients with delayed treatment intensification for more than two years. This difference could be because the reference study only involved those patients who need insulin treatment intensifications and also differences in healthcare systems[26].

The study’s strength is that, to the best of our knowledge, it is the first in the country to look at therapeutic inertia and how it affects diabetic treatment outcomes, by focusing on therapeutic inertia and its effect on the outcomes of diabetic treatment. This study can provide information for healthcare professionals that could guide them on where diabetes care needs to be improved. As a limitation, because our study is retrospective and depends only on available data, it was challenging to control for all variables, despite efforts have been made to account for confounding variables. We also used mean fasting blood sugar instead of HbA1c, which is a well-established measure of consistent glycaemia over time.

In conclusion, the findings of this study showed that diabetic patients exposed to therapeutic inertia had worse treatment outcome than non-exposed individual’s. Majority of general practitioners were major contributors to therapeutic inertia, indicating the need for more focused interventions and better training. Diabetic complications like nephropathy and neuropathy showed significant associations with therapeutic inertia when compared to non-inertia group. Therapeutic inertia was significantly associated with poor treatment outcome.

Based on the results of our study we would like to note the following recommendations. Wolaita Sodo University Comprehensive Specialized Hospital and MOH should implement training programs for general practitioners to improve their practices regarding timely treatment intensifications for type 2 diabetes patients. The hospital’s diabetic care clinic should promote the benefit of early glycemic control in order to reduce diabetic complications and poor treatment outcomes. The Hospital should give due attention for early glycemic control, so that poor diabetic outcome can be minimize. For researchers, prospective studies are needed to identify factors contributing to therapeutic inertia and also ongoing nationwide researches are required for a comprehensive strategy.

## Data Availability

Data cannot be shared publicly because of patient confidentiality and ethical restrictions. Data are available from the corresponding author (contact) for researchers who meet the criteria for access to confidential data.

## Author contributions

**Conceptualizations:** Dawit Alemu Lemma, Hailu chare Koyra.

**Data curation:** Dawit Alemu Lemma

**Formal Analysis:** Dawit Alemu Lemma.

**Investigations:**Dawit Alemu Lemma.

**Methodology:** Dawit Alemu Lemma, Hailu chare Koyra, Lijalem Abera Tema, Muluken Birhanu Mena.

**Writing - original draft:** Dawit Alemu Lemma, Hailu Chare Koyra, Lijalem Abera Tema.

**Writing - review and editing:** Dawit Alemu Lemma, Hailu Chare Koyra.

